# Greater Large Conducting Airway Luminal Area in Patients with Interstitial Lung Disease

**DOI:** 10.1101/2025.04.04.25324181

**Authors:** Alex J. Miller, Erik A. Ovrom, Solomiia Zaremba, Jonathon W. Senefeld, Chad C. Wiggins, Paolo B. Dominelli, Juan G. Ripoll, Brian T. Welch, Michael J. Joyner, Andrew H. Ramsook

**Affiliations:** Department of Anesthesiology and Perioperative Medicine, Mayo Clinic, Rochester, Minnesota, USA; Alix School of Medicine, Mayo Clinic, Rochester, Minnesota, USA; Department of Physiology and Biomedical Engineering, Mayo Clinic, Rochester, Minnesota, USA; Department of Health and Kinesiology, University of Illinois at Urbana-Champaign, Urbana, Illinois, USA; Beckman Institute for Advanced Science and Technology, University of Illinois at Urbana-Champaign, Urbana, Illinois, USA; Department of Kinesiology, Michigan State University, East Lansing, Michigan, USA; Department of Kinesiology and Health Sciences, University of Waterloo, Waterloo, Ontario, Canada; Department of Radiology, Mayo Clinic, Rochester, Minnesota, USA

**Keywords:** pulmonary fibrosis, dysanapsis, airway remodeling, CT imaging

## Abstract

Interstitial lung disease (ILD) encompasses multiple pulmonary disorders characterized by damaged pulmonary tissue caused by a sequence of inflammation and fibrosis. While much is known about ILD-associated changes within the parenchyma and the pathophysiological processes underpinning the disruption of pulmonary dynamics and gas exchange, less is known about ILD-associated changes of luminal area within the large conducting airways. We aimed to investigate whether luminal area of the large conducting airways is different between patients with ILD and healthy controls. In a retrospective case-control study, luminal areas of seven large conducting airways were measured using three-dimensional reconstructions of computed tomography imaging. Patients with ILD (N=82; 54% female) were compared to control subjects matched for age, sex, and height. Univariate ANOVA tests or Kruskal-Wallis tests were used to analyze group and sex differences. Patients with ILD had greater large conducting airway luminal areas than control subjects for measured large conducting airways, including the trachea (296±73 vs. 247±65 mm^2^, *P*<0.001), right main bronchus (214±59 vs. 161±46 mm^2^, *P*<0.001), bronchus intermediate (123±32 vs. 94±28 mm^2^, *P*<0.001), right upper lobe (81±22 vs. 61±20 mm^2^, *P*<0.001), left main bronchus (151±41 vs. 119±35 mm^2^, *P*<0.001), left lower lobe (71±22 vs. 48±15 mm^2^, *P*<0.001), and left upper lobe (86±21 vs. 68±22 mm^2^, *P*<0.001). Among patients with ILD, males had 17-34% greater luminal areas (normalized to height) than females depending on the airway segment (all *P*<0.05).

**NEW & NOTEWORTHY:** This study provides evidence that interstitial lung disease (ILD) is associated with greater large conducting airway luminal area, even if matched for key characteristics (age, sex, and height). Consistent with observations in health, adult males with ILD had greater height-normalized large conducting luminal areas than adult females with ILD.

## INTRODUCTION

Interstitial lung disease (ILD) encompasses a family of restrictive lung diseases characterized by chronic inflammation and resultant fibrosis (1, 2). ILD is associated with damage to the interstitial bed and alveoli which synergistically disrupts pulmonary gas exchange (3, 4). While pulmonary fibrosis in the lung interstitium of patients with ILD is well documented (5–8), less is known about how this disease may affect conducting airway luminal area.

Previous investigations examining the effect of ILD on airway luminal area have primarily focused on small (7-17 generations) airways (9–11). The small airways, coined the quiet zone (12), are conceptually considered a respiratory region where ILD may progress without detectable clinical signs and symptoms. Studies have indicated that patients with ILD have larger small airway luminal area and the development of honeycomb cysts in lungs with idiopathic pulmonary fibrosis (11). Many of these studies have been limited by using preclinical, rodent models of ILD, explanted human pulmonary tissue, or clinical studies with small sample sizes. Limited investigations have been performed examining the impact of ILD on *in vivo* human large conducting airways. In this framework, these large conducting airway luminal area measurements may provide a more comprehensive understanding of the pathophysiological effects associated with ILD.

Accordingly, the primary objective of our study was to determine the relationship between ILD and airway luminal area by comparing *in vivo* large conducting airway luminal area between patients with ILD and control subjects. This retrospective, case-control study used chest computed tomography (CT) scans to test the hypothesis that patients with ILD would have greater conducting airway luminal area than healthy control subjects matched for age, sex, and height. Additionally, because there are well-established sex differences in airway luminal area across the lifespan (13, 14) and the broader importance of considering sex as a biological variable (15), we included data for both males and females to assess potential sex differences in airway luminal area among patients with ILD.

## MATERIALS AND METHODS

### Ethical Approval

This retrospective study was approved by the Mayo Clinic Institutional Review Board (IRB #17-008537) and adhered to the standards set forth in the *Declaration of Helsinki*, except registration in a database. Routine clinical care resulted in CT scans of the thorax. Informed consent was waived as no identifiers were used in analyses, these clinical data already existed, the research did not affect patient care, and neither the patients nor their legal guardians opted out of their data being used for research. Consent waiver was approved by the Mayo Clinic Institutional Review Board.

### Patients

#### ILD patients

Initial inclusion criteria for ILD patients were 1) diagnosis by a pulmonologist, 2) at least one thoracic CT scan following their diagnosis, and 3) pulmonary function test (PFT) data. Exclusion criteria for patients included: age less than 18 years of age, any previous surgical intervention to the lungs, end stage liver disease, neoplasm, Sjögren’s syndrome, and additional cardiopulmonary disease (e.g., chronic obstructive pulmonary disease, asthma, heart failure, pleural effusion, obstructive sleep apnea, acute coronary syndrome, pulmonary infection). Listed conditions were excluded as they may have confounding effects on airway anatomy and pulmonary function. Exclusion criteria for CT scans included: poor scan resolution and incompatibility with the software used to measure airway luminal area. CT scans were selected on the basis of 1) scan resolution 2) temporal proximity between CT scan date and PFT date. CT scans were analyzed for large conducting airway cross-sectional luminal area.

#### Control subjects

Initial inclusion criterion was a chest CT scan for a suspected pulmonary embolism in adult participants over 18 years of age. In all cases, patients were negative for a pulmonary embolism. Similar exclusion criteria as used for ILD patients were applied. Additional exclusion criteria for control subjects included: ILD, idiopathic pulmonary fibrosis, tobacco use, end-stage kidney disease, ascites, and class III obesity (body mass index ≥ 40 kg·m^−2^) as these conditions may have confounding effects on airway anatomy and pulmonary function.

#### Patient matching

Using a nearest neighbor matching algorithm, potential control subjects and ILD patients were sequentially, individually matched (1:1) for sex, age, and height (**E-Figure 1**).

**Figure 1.**
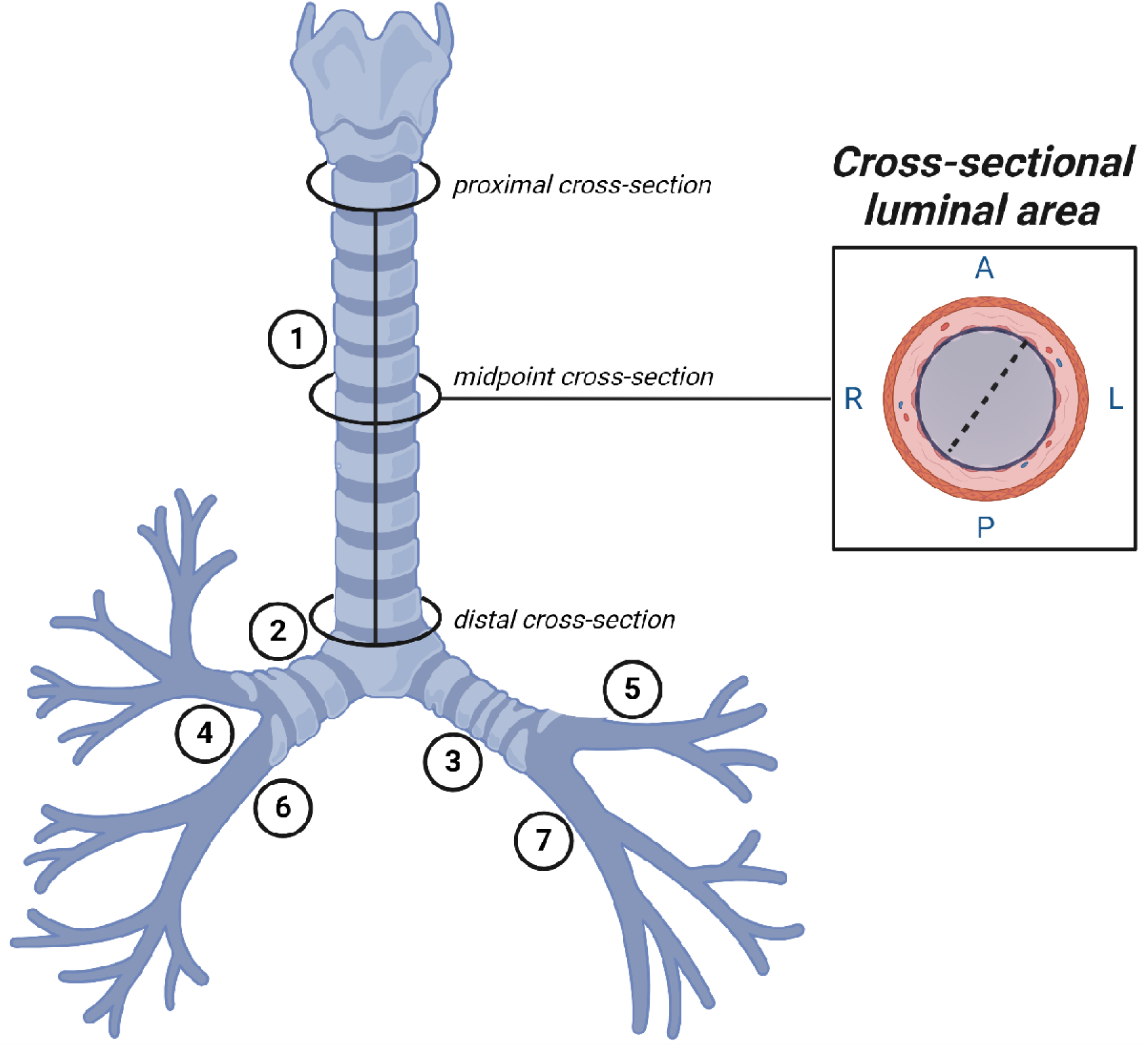
Graphical representation of the large conducting airway tree. Circles represent specific locations at which the cross-sections of the trachea (1) were taken. Other numbers represent additional airway segments measured. The dotted line within the inset represents a diameter measurement. 2, right main bronchus; 3, left main bronchus; 4, right upper lobe; 5, left upper lobe; 6, bronchus intermediate; 7, left lower lobe; A, anterior; L, left; P, posterior; R, right. Created with BioRender.com.

### Image acquisition

As described previously (13, 14, 16, 17), institutionally standardized CT algorithms are used for chest imaging at local medical centers. A posterior-anterior and lateral topogram is obtained at 120 kV and 35 mA. Spiral acquisitions with a pitch of 1.2 are utilized. Kilovoltage is set at 120 with a standard milliampere-second value of 140. Images are acquired at end-inspiration. Post imaging reconstructions are obtained in the axial and coronal plane using a B46 kernel. Slice thicknesses of 1.5 mm and 3.0 mm are reconstructed. Maximal intensity projections in the axial and coronal planes are completed with a slice thickness of 10 mm and reconstruction increment of 2.5 mm. participants inspired before imaging was performed. Because participants are not instructed to inhale maximally to total lung capacity, we were unable to match lung volumes (see **Limitations** below). Using image analysis, we measured lung volume and expressed it as a percent of total lung capacity for patients with ILD and as a percent of predicted total lung capacity for control subjects.

### Airway Measurements

Images were analyzed using commercially available software (TeraRecon, AQI, Foster City, CA, USA) as described previously (13, 14, 16, 17). After visual isolation from surrounding tissue, the software created a three-dimensional reconstruction from CT scans of the lungs and the airways. The three-dimensional reconstruction and optimized visualization enabled precision estimation of both absolute lung volume and cross-sectional luminal area of the large conducting airways. Airway measurements were all performed by single individual (AJM). A subsection of measurements were validated against independent training samples used in the laboratory and repeated by multiple investigators (SZ and AHR). The cross-sectional areas of conducting airway segments were digitally measured at three points (proximal, middle, and distal points) for each of the following airway segments: the trachea, right and left main bronchus, right and left upper lobes, bronchus intermediate, and left lower lobe. The proximal cross-section of the trachea was defined as the point below the cricoid cartilage. The distal cross-section was defined as the point above its bifurcations. Anatomical bifurcations defined the proximal and distal points of the measured bronchi. The midpoint was defined as half the length of the airway segment. A graphical representation of an airway tree describing cross-sectional area measurements is displayed in **Figure 1**. For the analysis of sex differences in ILD patients, airway luminal areas were represented relative to patient height (“height-normalized”) using the quotient of airway luminal area (mm^2^) to patient height (cm).

### Pulmonary Function Testing (PFT)

Institutional standards of clinical care for patients with ILD include routine PFTs. Thus, when available, PFT data (total lung capacity [TLC], volume/TLC, ratio of forced expired volume in one second to forced vital capacity [FEV_1_/FVC], and diffusing capacity for carbon monoxide [D_LCO_]) were abstracted from medical records of patients with ILD. CT scans were selected to optimize temporal proximity relative to PFT collection. PFT data were not available in control cohort thus predicted values were used for the purpose of comparison.

### Statistical analysis

Normality was assessed using Shapiro-Wilk tests and homoscedasticity was assessed using Levene’s test. Statistical models were determined based on normality and homoscedasticity. Univariate analyses of variance (ANOVA) were used to compare participant characteristics (age, height, mass, body mass index, days since diagnosis, and smoking history) and pulmonary characteristics (computed lung volume, TLC, volume/TLC, FEV_1_/FVC, and D_LCO_) between patients with ILD and matched control subjects. ANOVAs, separated by airway segment, were used to compare metrics of conducting airway luminal area between patients with ILD and matched control subjects. One-way ANOVA statistical models were performed using a representation of airway luminal area— the measurement of the average of three cross-sectional areas (proximal, middle, and distal). Descriptive statistics are presented as mean ± standard deviation (SD) within the text and tables for normally distributed data and are presented as median (IQR) for non-normally distributed data. *P* values were reported, and the interpretation of findings was based on *P*<0.05. Analyses were performed with the use of IBM Statistical Package for Social Sciences version 28 statistical package (Armonk, New York, USA).

Notably, several distributions of large conducting airway luminal cross-sectional area failed tests of normality. Tukey’s fences were used to identify statistical outliers. Tukey’s methods identify values that are 1.5 IQR below Q1 and 1.5 IQR above Q3 as outliers, where IQR denotes interquartile range, Q1 denotes lower quartile, and Q3 denotes upper quartile. In this context, outliers approximate the 1st or 99th percentile of a dataset. After removal of outliers, tests of normality were performed again. If sample distributions failed tests of normality after removal of outliers, Kruskal-Wallis tests were used. If tests of normality were passed after the removal of outliers, univariate ANOVA tests were performed on the dataset sans outliers. Among males, five airways were normally distributed (trachea, right main bronchus, bronchus intermediate, right upper lobe, and lower main bronchus) and two airways failed tests of normality (left lower lobe and left upper lobe). However, after removing outliers the left upper lobe no longer failed tests of normality thus parametric tests were used for statistical analyses. The left lower lobe continued to fail tests of normality thus non-parametric tests were used for statistical analyses. Among females, three airways were normally distributed (bronchus intermediate, right upper lobe, and lower main bronchus) and four airways failed tests of normality (trachea, right main bronchus, left lower lobe and left upper lobe). However, after removing outliers, the trachea, right main bronchus, and left lower lobe no longer failed tests of normality. The left upper lobe continued to fail tests of normality thus non-parametric tests were used for statistical analyses.

## RESULTS

### Screening

#### ILD patients

Medical histories of 804 patients with ILD who met the initial search criteria were screened by the research team. After screening, CT scans of 163 patients with ILD met the inclusion criteria were screen for potential assessment of airway luminal area. During data abstraction, 81 additional patients with ILD were excluded due to incomplete PFT data (N = 72), CT scans performed before ILD diagnosis (N = 7), or poor CT scan quality (defined by inability to aptly visualize one or more of the large conducting airways, N = 2). The final cohort of patients with ILD analyzed in this study represented 82 patients, including 38 males and 44 females.

#### Control subjects and patient matching

Medical histories of 2,034 participants who met the initial search criteria were screened. After exclusion, 132 participants were considered for matching and their CT scans were analyzed for large conducting cross-sectional luminal area. Using our nearest neighbor matching algorithm, another 50 participants were excluded due to inadequate demographic and anthropometric characteristic for matching.

For the control cohort, 132 participants (51 males, 81 females) who were included in our previous study (14) were considered for inclusion. Using a nearest neighbor matching algorithm, potential control subjects were individually (1:1) matched for sex, age, and height (**E-Figure 1**). In this framework, the final cohort of control subjects was analogous to the ILD patients and included 82 participants (38 males and 44 females). The participants inclusion paradigm is displayed in **Figure 2**.

**Figure 2.**
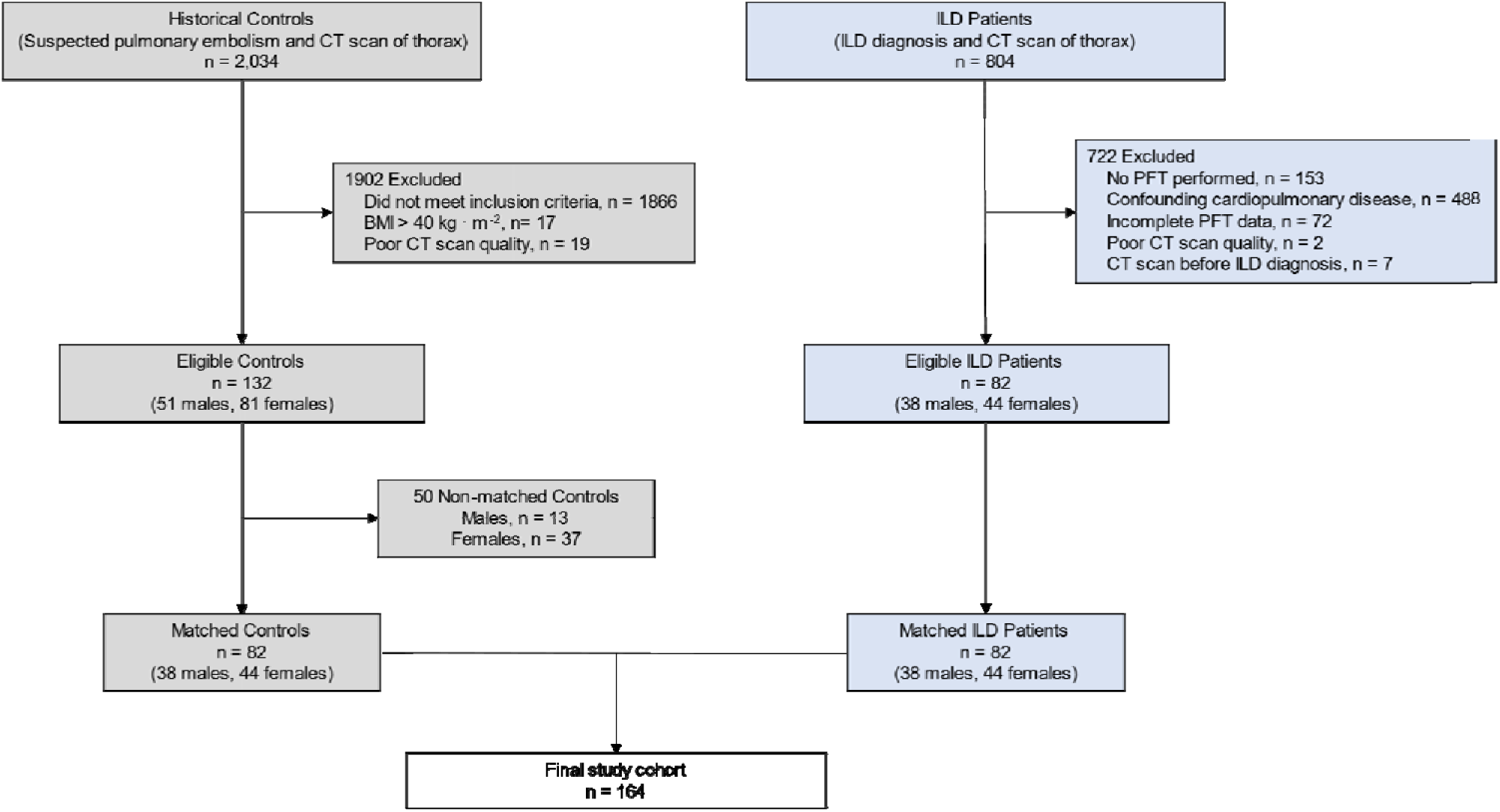
Participant inclusion paradigm of patient eligibility and control subject matching for the study. Abbreviations: BMI, body mass index; CT, computer tomography; ILD, interstitial lung disease; PFT, pulmonary function test.

#### Patient characteristics

Patient characteristics are displayed in **Table 1**. Control subjects had a lower computed scan volume relative to predicted TLC compared to patients with ILD (*P*<0.05). Among males, there were no group differences (ILD vs control subjects) in mass, body mass index (BMI), and computed volume. However, females with ILD had smaller mass and BMI compared to female control subjects (*P*<0.05). Females with ILD also had larger computed lung volume (*P*<0.05) compared to female control subjects. Subgroups of specific ILD diagnoses are presented, **E-Table 2**.

**Table 1.** Patient characteristics.

| Variable | Males |  |  | Females |  |  |
| --- | --- | --- | --- | --- | --- | --- |
|  | ILD | Control | P-value | ILD | Control | P-value |
| Cohort Size, *n* | 38 | 38 | - | 44 | 44 | - |
| *Participant Characteristics* |  |  |  |  |  |  |
| Age, years | 62 ± 10 | 60 ± 13 | 0.459 | 61 ± 14 | 61 ± 14 | 0.796 |
| Height, cm | 177 ± 5 | 178 ± 7 | 0.350 | 162 ± 7 | 163 ± 6 | 0.379 |
| Weight, kg | 92 ± 15 | 95 ± 14 | 0.383 | 74 ± 17 | 90 ± 24 | **<0.001** |
| BMI, kg·m<sup>-2</sup> | 30 ± 4 | 30 ± 5 | 0.537 | 28 ± 6 | 34 ± 9 | **<0.001** |
| Days Since Diagnosis | 495 ± 1003 | - | - | 366 ± 801 | - | - |
| Smoking History, pack/years | 23 ± 30 | - | - | 7 ± 15 | - | - |
| *Scan and PFT* |  |  |  |  |  |  |
| Computed Volume, cm<sup>3</sup> | 4106 ± 1331 | 4171 ± 1548 | 0.848 | 3266 ± 1030 | 2822 ± 729 | **0.023** |
| Computed Volume/TLC, % | 85 ± 17 | 58 ± 21 | **<0.001** | 83 ± 18 | 55 ± 13 | **<0.001** |
| TLC, liters | 4.7 ± 1.6 | 7.3 ± 0.7<sup>†</sup> | **<0.001** | 3.8 ± 1.0 | 5.2 ± 0.4<sup>†</sup> | **<0.001** |
| TLC, % predicted | 71 ± 21 | - | - | 70 ± 19 | - | - |
| FEV<sub>1</sub>/FVC | 0.79 ± 0.1 | 0.78 ± 0.02<sup>†</sup> | 0.244 | 0.77 ± 0.1 | 0.79 ± 0.02<sup>†</sup> | 0.291 |
| FEV<sub>1</sub>/FVC, % predicted | 101 ± 9 | - | - | 97 ± 17 | - | - |
| D<sub>LCO</sub>, mL<sub>CO</sub>/sec/mmHg | 15 ± 7 | - | - | 12 ± 4 | - | - |
| D<sub>LCO</sub>, % predicted | 61 ± 26 | - | - | 57 ± 17 | - | - |

Data are reported as count or mean ± standard deviation (SD). Data are compared using separate univariate ANOVAs. *P*-values are reported for between group comparisons (ILD vs. control subjects) for males and females separately. Abbreviations: BMI, body mass index TLC, total lung capacity; FEV<sub>1</sub>/FVC, forced expiratory volume in the first one second over forced vital capacity; D<sub>LCO</sub>, diffusing capacity of the lungs for carbon monoxide. “-” indicates no data available. “†” indicates predicted values used as no other data was available.

**Table 2.**
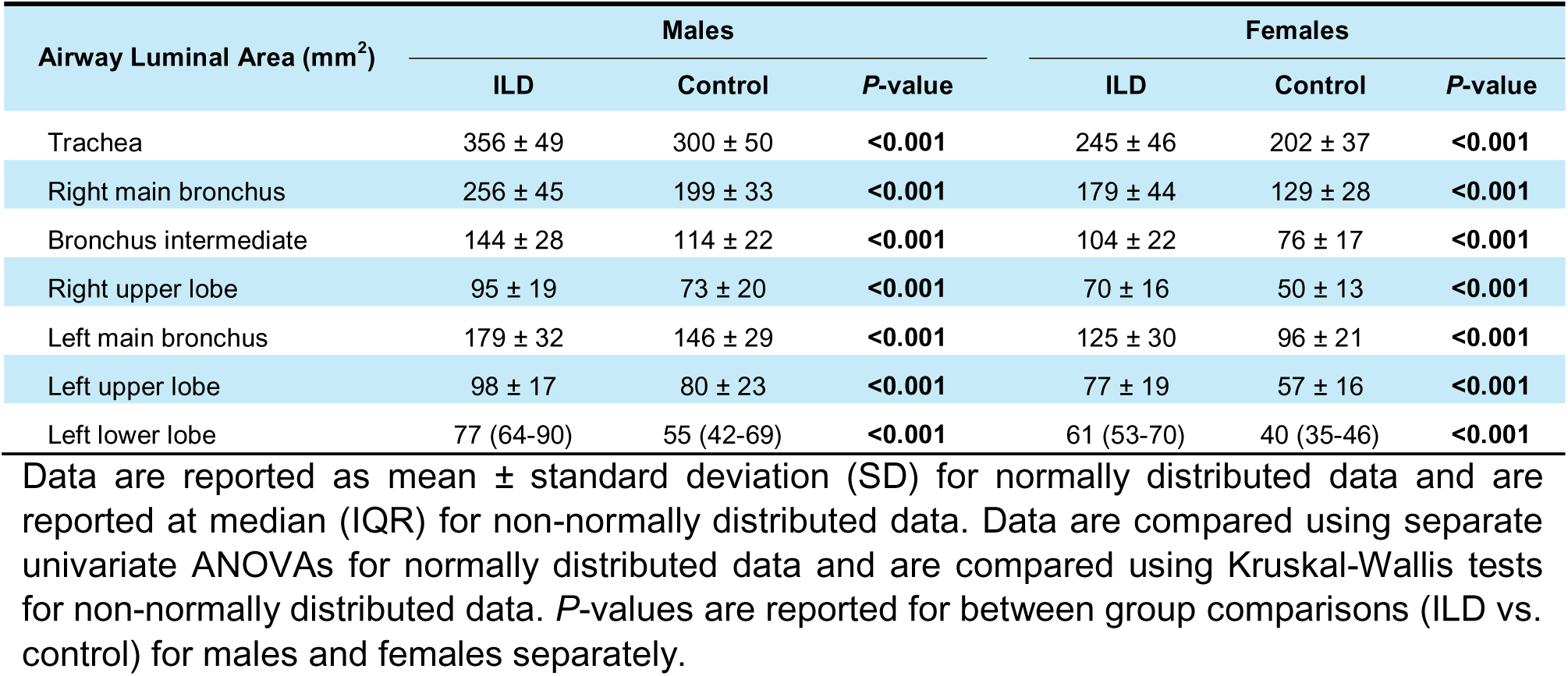
Airway luminal area of males and females diagnosed with ILD and a height- and age-matched control cohort.

| Airway Luminal Area (mm<sup>2</sup>) | Males |  |  | Females |  |  |
| --- | --- | --- | --- | --- | --- | --- |
|  | ILD | Control | *P*-value | ILD | Control | *P*-value |
| Trachea | 356 ± 49 | 300 ± 50 | <0.001 | 245 ± 46 | 202 ± 37 | <0.001 |
| Right main bronchus | 256 ± 45 | 199 ± 33 | <0.001 | 179 ± 44 | 129 ± 28 | <0.001 |
| Bronchus intermediate | 144 ± 28 | 114 ± 22 | <0.001 | 104 ± 22 | 76 ± 17 | <0.001 |
| Right upper lobe | 95 ± 19 | 73 ± 20 | <0.001 | 70 ± 16 | 50 ± 13 | <0.001 |
| Left main bronchus | 179 ± 32 | 146 ± 29 | <0.001 | 125 ± 30 | 96 ± 21 | <0.001 |
| Left upper lobe | 98 ± 17 | 80 ± 23 | <0.001 | 77 ± 19 | 57 ± 16 | <0.001 |
| Left lower lobe | 77 (64-90) | 55 (42-69) | <0.001 | 61 (53-70) | 40 (35-46) | <0.001 |

Data are reported as mean ± standard deviation (SD) for normally distributed data and are reported at median (IQR) for non-normally distributed data. Data are compared using separate univariate ANOVAs for normally distributed data and are compared using Kruskal-Wallis tests for non-normally distributed data. *P*-values are reported for between group comparisons (ILD vs. control) for males and females separately.

### Airway luminal area: ILD vs. Health

In all measured large conducting airways, average luminal area was greater among patients with ILD compared to control subjects, (**Figure 3**; all *P*<0.001). Depending on the airway segment, cross-sectional luminal area was 20-48% greater in ILD patients compared to control subjects, **Table 2**.

**Figure 3.**
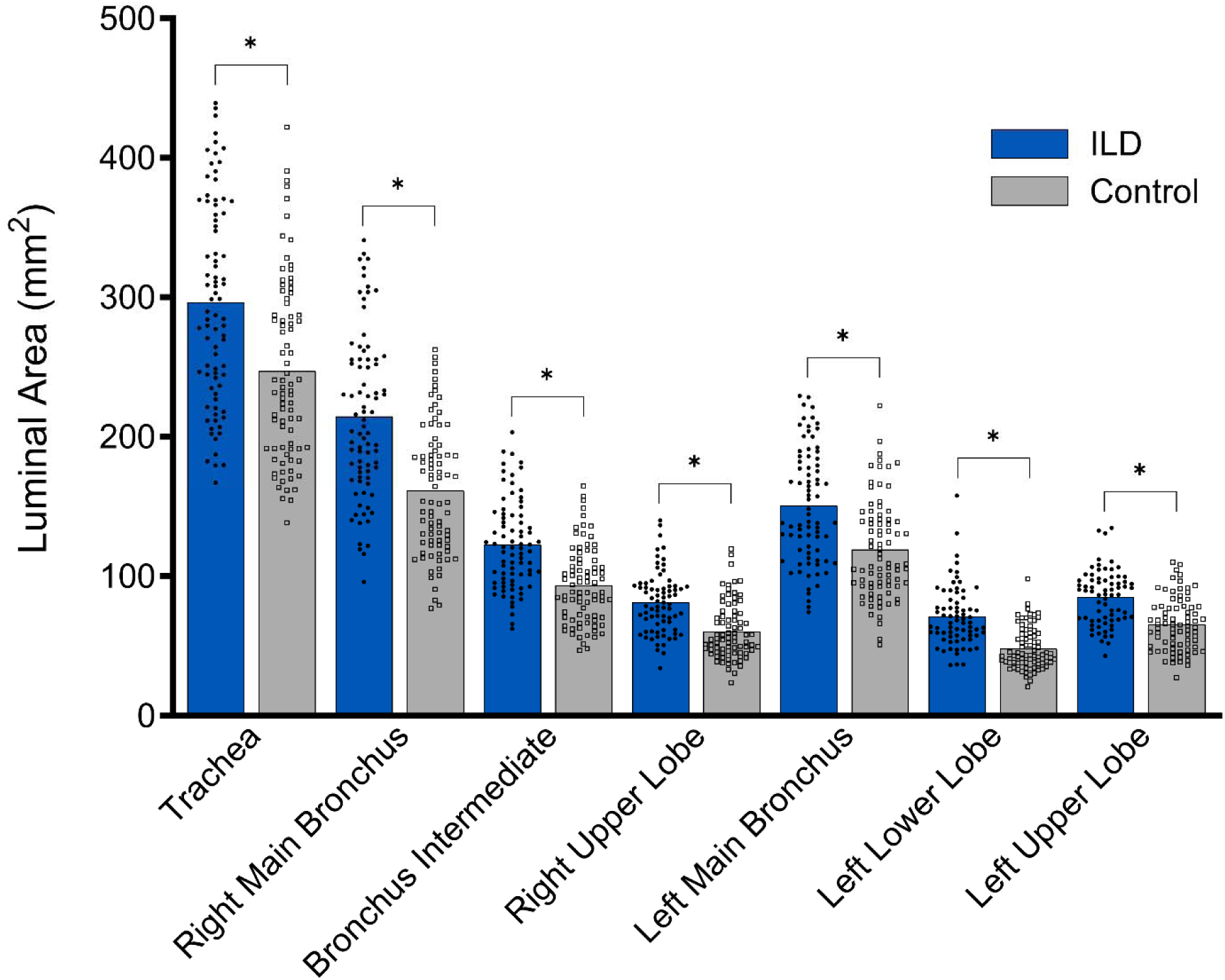
Luminal airway area of patients with ILD and healthy, sex-, height-, and age-matched control subjects. *, *P*<0.05 indicating significant group difference. Data are presented as mean with individual data. Outlier data are excluded.

### Sex-related differences in ILD

In all measured large conducting airways, height normalized luminal area was greater in males with ILD compared to females with ILD, **Figure 4** (*P*<0.001). Height normalized luminal area of males with ILD was 19-34% greater depending on the airway segment than females with ILD, **Table 3**.

**Figure 4.**
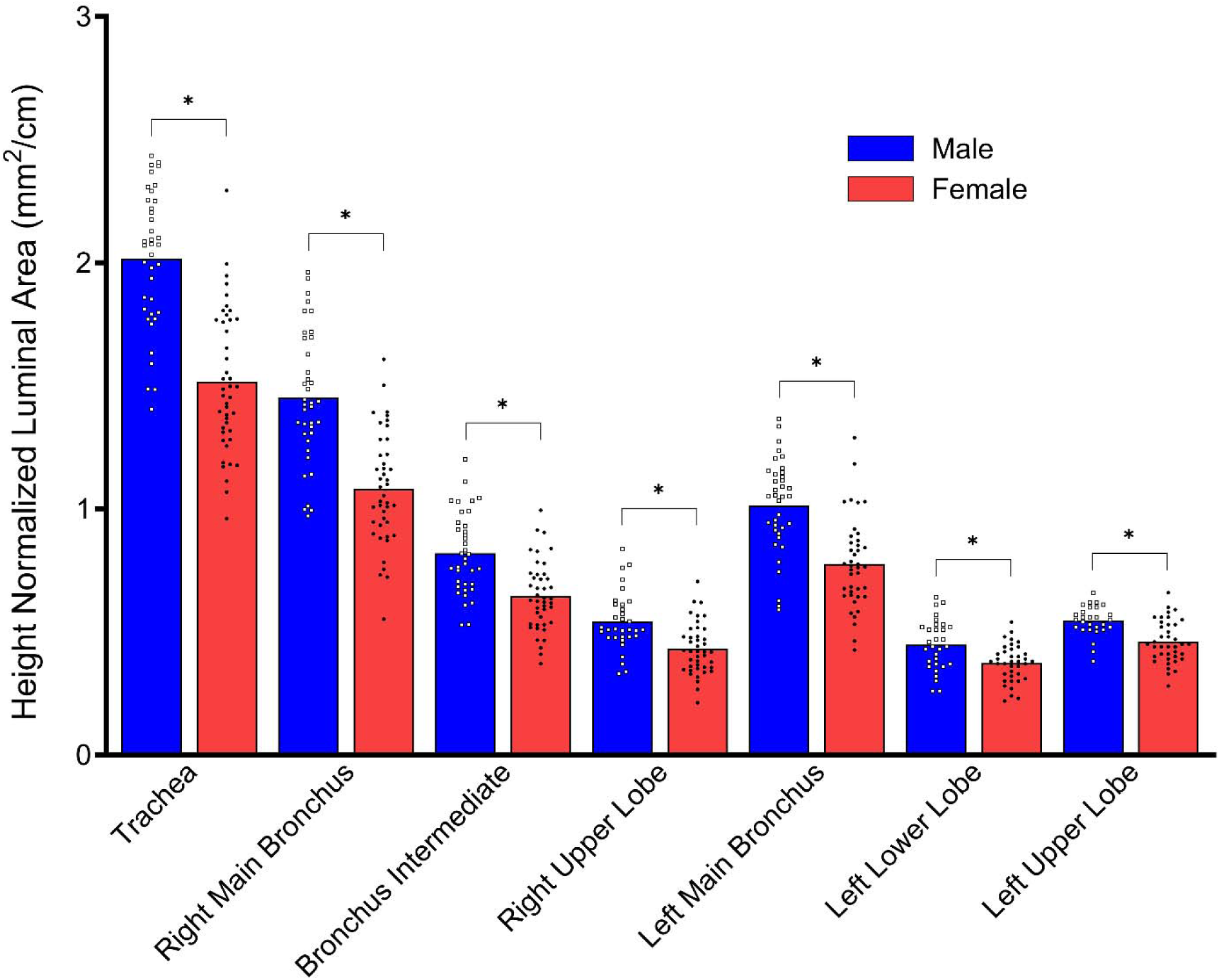
Luminal airway area normalized to height in males and females with ILD. *, *P*<0.05 indicating significant biological sex difference. Data are presented as mean with individual data. Outlier data are excluded.

**Table 3.** Airway luminal area (mm^2^) normalized to height (cm) of males and females diagnosed with ILD.

| Height Normalized Airway Luminal Area (mm<sup>2</sup>/cm) | ILD |  |  |
| --- | --- | --- | --- |
|  | Males | Females | *P*-value |
| Trachea | 2.02 ± 0.28 | 1.51 ± 0.29 | <0.001 |
| Right main bronchus | 1.45 ± 0.27 | 1.08 ± 0.23 | <0.001 |
| Bronchus intermediate | 0.82 ± 0.16 | 0.64 ± 0.14 | <0.001 |
| Right upper lobe | 0.54 ± 0.12 | 0.43 ± 0.10 | <0.001 |
| Left main bronchus | 1.01 ± 0.19 | 0.78 ± 0.18 | <0.001 |
| Left upper lobe | 0.54 ± 0.06 | 0.46 ± 0.09 | <0.001 |
| Left lower lobe | 0.45 ± 0.10 | 0.37 ± 0.08 | <0.001 |

Data are reported as mean ± standard deviation (SD). Data are compared using separate univariate ANOVAs. *P*-values are reported for intra-group sex differences (males with ILD vs. females with ILD).

## DISCUSSION

The primary aim of this study was to evaluate large conducting airway luminal area among patients with ILD compared to control subjects matched for age, sex, and height. In support of our primary hypothesis, we found that patients with ILD had greater cross-sectional luminal areas compared to matched control subjects. Exploratory analyses demonstrated that the expected sex difference persisted in patients with ILD, resulting in males having greater cross-sectional luminal areas in the large conducting airways than females. Notably, this observed sex difference in large conducting airway luminal area is also commonly observed in healthy youths (13) and adults (14, 18).

### Pathophysiology of ILD in the conducting airways

Previous research has focused on the effects of fibrosis on surfaces of the lungs responsible for gas exchange (19–22). Findings suggest that ILD is associated with an increase in diameter of small airways (10, 11). As the lungs become inflamed and consequently fibrosed throughout the progression of ILD, compliance (23–25) decreases thereby causing a restrictive breathing pattern and ineffective ventilation (26–28). In this context, a decrease in ventilation due airway dilation and a resultant increase in anatomic dead space (29) may compound respiratory dysfunction.

One potential explanation for our findings is progressive fibrosis extending from the parenchyma into the conducting airways. Ikezoe et al. investigated fibrosis in the small conducting airways and discovered concomitant increases in markers of fibrosis and increases in luminal area (11). Pulmonary fibrosis in the large conducting airways leads to reduced compliance (23) and elasticity rendering the conducting airways dilated and rigid. Coupling this pathophysiological process with concomitant decreases in compliance of the surrounding pulmonary tissue (24) leads to an increased metabolic cost of breathing (30). This framework is supported by our findings as airway luminal area was greater in patients with ILD, which may indicate fibrotic tissue in the large conducting airways and may help explain increased work of breathing (31), found in an animal model, and dyspnea in ILD (32, 33). Overall, in the context of previous studies, our findings suggest that ILD disrupts the lungs more extensively than what has been thought to be the disease’s primary target, the bronchovascular interstitium.

### Sex differences in airway anatomy

We found that among patients with ILD, males had greater luminal areas normalized to height in the large conducting airways than females. Limited research has been performed to examine potential biological sex differences in airway diameter among patients with ILD (10, 11, 34–36), although sex differences in airway luminal area are commonly observed in healthy control subjects across the lifespan, even when adjusted for factors that commonly affect airway size, like height. Epidemiological data has shown that males with ILD have lower survival rates, increased prevalence of disease, and attenuated lung function (37–39). Such findings warrant investigation into the role of sex specific hormones in pulmonary fibrosis however the effect of biological sex hormones on pulmonary fibrosis and airway remodeling remains unclear (40–43). Previous studies examining sex differences in airway luminal area among patients with ILD have been limited to mice models, using *ex vivo* human lungs, lacking a comparator control group, and/or evaluating only the small airways. Our research aimed to address some of these gaps in the literature.

### Potential clinical implications

As pulmonary fibrosis progresses, compliance of the conducting airways and parenchyma are reduced (44). Thus, lung volumes are decreased. In this framework, pulmonary fibrosis in the interstitial space results in a compounding effect of conducting airway dysfunction and a reduction in gas exchanging tissue leading to an even greater reduction of the diffusive capabilities of the lungs.

Future work should investigate the mechanisms behind our findings of greater large conducing airway luminal area. Characterizing potential histopathological changes in the large conducting airways may explain our findings of increased luminal area. If markers of fibrosis and inflammation are found in the large conducting airways of patients with ILD, comprehensive treatment of ILD might include interventions to not only target the sequalae of inflammation and fibrosis in the parenchyma but also dysfunction and remodeling within the conducting airways

(45). However, in order to systematically treat the downstream effects of ILD such as dyspnea, we must comprehensively understand the effect of the disease not only in the parenchyma but also the small and large conducting airways.

### Limitations

Several limitations resulted from the design of this retrospective study. First, the end-inspiratory lung volume was not standardized to total lung capacity as participants were only instructed to inspire and hold their breath as many scans were performed under emergency circumstances (*e.g.*, pulmonary embolism). Lung volume is likely to have the greatest influence on more distal airway compared to first and second generation airways (46). Second, our methodology and study design only allow for observational conclusions. Our findings suggest an association between ILD and greater airway luminal area but do not infer a causal relationship. Third, PFT data were not available for healthy control subjects and predicted values were calculated and used for the purpose of group comparisons.

### Conclusions

These data provide evidence that patients with ILD have significantly greater cross-sectional airway luminal area than matched control subjects. We also conclude that males with ILD have significantly greater cross-sectional airway area normalized to height than females with ILD suggesting absent sex differences in large conducting airway luminal area changes. Overall, these findings may, in part, explain symptomatology of ILD such as dyspnea. This work warrants future investigation into the mechanisms behind our finding of increased large conducting airway luminal area. Discovery of novel components of the pathophysiology of ILD may provide potential targets for pharmaceutical and medical intervention and as a result improve clinical outcomes and morbidity in patients with ILD.

## DATA AVAILABILITY

Study data cannot be shared publicly because the Institutional Review Board restrictions. Individual participant data underlying the results reported in this publication may be made available to approved investigators for secondary analyses. A scientific committee will review requests for the conduct of protocols approved or determined to be exempt by an Institutional Review Board. Requestors may be required to sign a data use agreement. Data sharing must be compliant with all applicable Mayo Clinic policies.

## ACKNOWLEDGMENTS

Graphical abstract and Figure 1 created with BioRender and published with permission.

Present addresses: E. A. Ovrom, University of Chicago, Chicago, Illinois; P. B. Dominelli, University of Calgary, Calgary, Alberta, Canada.

## AUTHOR CONTRIBUTIONS

A.J.M., J.W.S., C.C.W., and A.H.R. conceived and designed the research; A.J.M., E.A.O., and A.H.R. performed experiments; A.J.M., J.W.S., C.C.W., and A.H.R. analyzed data; A.J.M., S.Z., J.W.S., C.C.W., and A.H.R. interpreted results of experiments; A.J.M. and S.Z. prepared figures; A.J.M., J.W.S., C.C.W., and A.H.R. drafted the manuscript; A.J.M., E.A.O., S.Z., J.W.S., C.C.W., P.B.D., J.G.R., B.T.W., M.J.J., and A.H.R. edited and revised the manuscript. A.J.M., E.A.O., S.Z., J.W.S., C.C.W., P.B.D., J.G.R., B.T.W., M.J.J., and A.H.R. approved the final version of the manuscript.

