## Supplemental Materials for "Greater Large Conducting Airway Luminal Area in Patients with Interstitial Lung Disease"

This appendix has been provided by the authors to give readers additional information about their work.

**Most Recent Update:** April 04, 2025

**Greater Large Conducting Airway Luminal Area in**

**Patients with Interstitial Lung Disease**

**Supplementary Appendix**

**Contents**

**E-Table 1.** Airway size of men and women previously diagnosed with interstitial lung disease (ILD) and a height- and age-matched control cohort.	3

**E-Table 2.** ILD diagnosis.	4

**E-Figure 1.** Matches chosen using 1:1 nearest neighbor matching algorithm based on height of males (dark blue squares) and females (red circles) with ILD and controls.	5

**E-Table 1.** Airway size of men and women previously diagnosed with interstitial lung disease (ILD) and a height- and age-matched control cohort. Airway size data are the midpoints of each airway. Data are reported as mean ± standard deviation (SD) for normally distributed data and are reported at median (IQR) for non-normally distributed data. Data are compared using separate univariate ANOVAs for normally distributed data and are compared using Kruskal-Wallis tests for non-normally distributed data. *P*-values are reported for between group comparisons (ILD vs. control) for men and women separately.

| **Airway Luminal Size** | **Men** | | |  | **Women** | | |
| --- | --- | --- | --- | --- | --- | --- | --- |
|  | **ILD** | **Control** | ***P*-value** |  | **ILD** | **Control** | ***P*-value** |
| Trachea, mm^2^ | 353 ± 68 | 294 ± 60 | **<0.001** |  | 250 ± 56 | 204 ± 42 | **<0.001** |
| Right main bronchus, mm^2^ | 251 ± 45 | 192 ± 32 | **<0.001** |  | 174 ± 47 | 125 ± 28 | **<0.001** |
| Right upper lobe, mm^2^ | 89 ± 19 | 71 ± 21 | **<0.001** |  | 64 ± 16 | 49 ± 13 | **<0.001** |
| Bronchus intermediate, mm^2^ | 136 ± 28 | 109 ± 24 | **<0.001** |  | 97 ± 23 | 70 ± 19 | **<0.001** |
| Left main bronchus, mm^2^ | 167 ± 34 | 128 ± 27 | **<0.001** |  | 112 ± 32 | 81 ± 21 | **<0.001** |
| Left upper lobe, mm^2^ | 96 (89-104) | 75 (59-91) | **0.001** |  | 74 ± 19 | 56 ± 16 | **<0.001** |
| Left lower lobe, mm^2^ | 65 (52-79) | 50 (37-63) | **<0.001** |  | 56 (49-64) | 39 (32-46) | **<0.001** |

**E-Table 2.** ILD diagnosis

| **Classification** | **N** |
| --- | --- |
| Idiopathic Interstitial Pneumonia | **31** |
| Idiopathic Pulmonary Fibrosis | **27** |
| Hypersensitivity Pneumonitis | **12** |
| CTD-associated ILD | **3** |
| Unclassified ILD | **3** |
| Idiopathic Lymphoid interstitial pneumonia | **2** |
| Scleroderma | **2** |
| Polymyositis-associated ILD | **1** |
| Granulomatous interstitial lung disease | **1** |

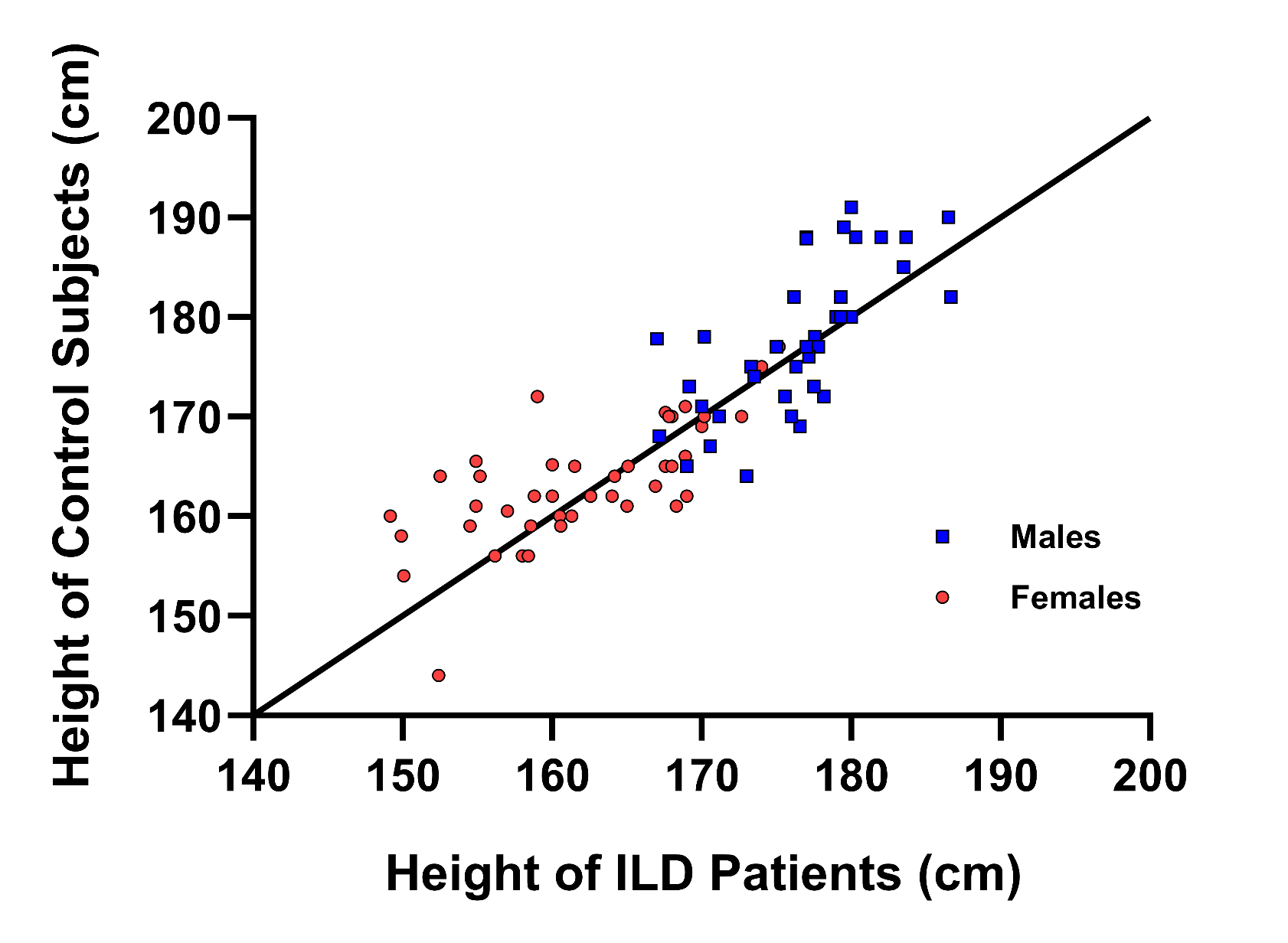
**E-Figure 1.** Matches chosen using 1:1 nearest neighbor matching algorithm based on height of males (dark blue squares) and females (red circles) with ILD and control subjects. Symbols represent height of patients with ILD and control subjects, and the line of identity represents optimal height matching.
